# Estimating the impact of COVID-19 vaccine allocation inequities: a modeling study

**DOI:** 10.1101/2022.11.18.22282514

**Authors:** Nicolò Gozzi, Matteo Chinazzi, Natalie E. Dean, Ira M. Longini, M. Elizabeth Halloran, Nicola Perra, Alessandro Vespignani

## Abstract

Access to COVID-19 vaccines on the global scale has been drastically impacted by structural socio-economic inequities. Here, we develop a data-driven, age-stratified epidemic model to evaluate the effects of COVID-19 vaccine inequities in twenty lower middle and low income countries (LMIC) sampled from all WHO regions. We focus on the first critical months of vaccine distribution and administration, exploring counterfactual scenarios where we assume the same per capita daily vaccination rate reported in selected high income countries. We estimate that, in this high vaccine availability scenario, more than 50% of deaths (min-max range: [56% − 99%]) that occurred in the analyzed countries could have been averted. We further consider a scenario where LMIC had similarly early access to vaccine doses as high income countries; even without increasing the number of doses, we estimate an important fraction of deaths (min-max range: [7% − 73%]) could have been averted. In the absence of equitable allocation, the model suggests that considerable additional non-pharmaceutical interventions would have been required to offset the lack of vaccines (min-max range: [15% − 75%]). Overall, our results quantify the negative impacts of vaccines inequities and call for amplified global efforts to provide better access to vaccine programs in low and lower middle income countries.

---

Throughout the COVID-19 pandemic, socio-economic disparities have been linked to higher and disproportionate COVID-19 burden, a finding replicated across different countries and social strata [1–10]. In this context, structural inequities in access to COVID-19 vaccines were discussed even before any specific vaccine was authorized by regulatory agencies [11–15]. Despite international initiatives for equitable sharing agreements such as the COVID-19 Global Vaccine Access (COVAX) program [16, 17], vaccine nationalism has largely superseded global equity efforts. Indeed, the differences in terms of COVID-19 vaccines doses, administered across countries grouped by income levels, are staggering [18–21]. These inequities have potentially enormous effects on the economies and future health of lower middle and low income countries (LMIC) [11, 12, 22–25]. However, except for three recent studies that modelled the effects of different global vaccine allocation and sharing strategies [26–28], a quantitative, detailed, and tailored estimation of the consequences of vaccine inequity across several LMIC is largely missing.

## Quantifying vaccine inequities

To quantify vaccine distribution and administration across countries with different income levels, we combine two datasets. The first one is collected and updated by the United Nations Development Programme via their Global Futures Platform [18]. It provides general information about COVID-19 vaccine data around the world, including several socio-economic dimensions. The second dataset, made available by Our World in Data [29], provides a range of general information about country-level vaccination efforts, including the number of doses administered as a function of time. As of October 1^*st*^ 2022, 77% of individuals that completed the initial COVID-19 vaccination course (i.e., one or two doses depending on the vaccine) are living in high and upper middle income countries. The equivalent share in LMIC was 50%, around 1.5 times lower. Inequities were drastically more pronounced during the earliest stages of the vaccination rollout, when populations also had much lower levels of infection-induced immunity. In Figure 1-A, we plot the total number of doses administered per 100 people. By October 1^*st*^, 2021 (i.e., the horizon of our analysis), high and upper middle income countries have, on average, more than one dose per person. The numbers are drastically different in LMIC. While lower middle income countries administered slightly more than 40 doses per 100, the equivalent number for low income countries is only 3.6 doses per 100.

**Figure 1:**
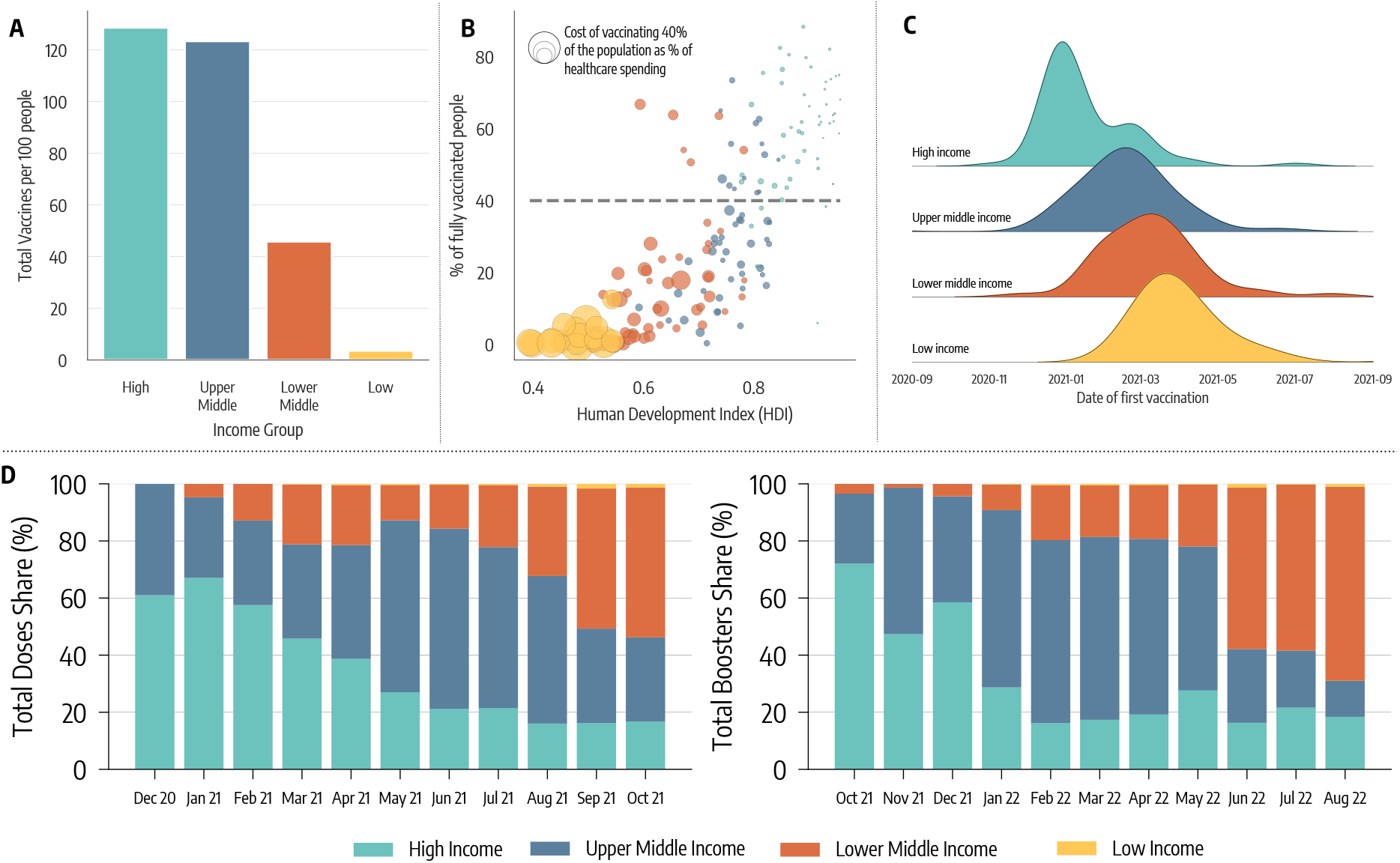
Vaccine inequities. A) Total number of doses administered per 100 people in different income groups as of October 1, 2021. B) Scatter plot of % of a country’s population who is fully vaccinated versus their Human Development Index (HDI). The color of dots indicates the country’s income group while size is proportional to the cost of vaccinating 40% of the population as a percentage of current healthcare spending. C) Density plot of the date of first COVID-19 vaccination across different country income groups. D) Evolution in the share of doses administered monthly across country income groups (left hand), and evolution of monthly booster doses share (right hand).

In Figure 1-B, we plot the percentage of each country’s population that was fully vaccinated by October 1^*st*^, 2021, versus the country’s Human Development Index (HDI). The HDI is a composite index that accounts for life expectancy, education, and per capita income as well as other aspects of human development [30]. The size of the data points is set proportional to the estimated cost of vaccinating 40% of the population as a percentage of the country’s current healthcare spending; this metric is used to quantify the economic challenge posed by achieving the 40% vaccination level proposed by WHO has an interim target by the end of 2021 [18]. The plot shows a strong positive correlation between HDI and vaccination coverage (Pearson correlation coefficient: 0.79, *p <* 0.001). The more developed the country, the higher the fraction of its population vaccinated. Furthermore, countries characterized by the lowest values of HDI face drastically bigger economic challenges in reaching 40% of the population vaccinated. Unfortunately, even one year later, as of October 1^*st*^ 2022, only half of lower middle income countries have vaccinated at least 40% of the population, while only 1 low income country out of 10 met the target.

In Figure 1-C, we plot the distribution of the vaccine administration starting dates by country income group. For high and upper middle income countries this date is generally much earlier than for LMIC. This figure clearly highlights the different logistic challenges in setting up a mass vaccination campaign across countries and speaks to the differential power among income levels to secure a scarce resource such as COVID-19 vaccines in the early phases of the rollout [19].

In Figure 1-D, we plot the share of vaccines administered globally across country income levels by month, starting in December 2020. The plot confirms that, in the first seven months of the COVID-19 vaccination campaign, more than 80% of the doses were concentrated in high and upper middle countries. The level of inequity becomes even more staggering when considering the share of the global population of the four groups. Only 16% of the global population lives in high income countries, while nearly 50% live in LMIC (8% in low income countries). Hence, the plot also highlights how, as of the 1^*st*^ of October 2021, low income countries had administered a share of doses that is smaller than 1% of total doses. As shown on the right hand of Figure 1-D, high income countries similarly monopolized the administration of booster doses in late 2021 and the first half of 2022, when most LMIC were still far behind with primary vaccinations.

## Modeling the vaccination campaigns in LMIC

To quantify the impact of vaccines inequities on COVID-19 burden, we developed a stochastic and multi-strain epidemic model. We consider an SLIR-like natural history of the disease, and the model takes as input for each country its demographics, non-pharmaceutical interventions (NPIs), age-stratified contact matrices from Ref. [31], variants prevalence, and epidemic data describing confirmed deaths. Vaccine administration is explicitly modeled with the number of daily 1^*st*^ and 2^*nd*^ doses in different countries from Ref. [32]. The model is stochastic and transitions among compartments are simulated through chain binomial processes. We consider individuals who received one or two vaccine doses, with vaccine efficacy against infection and death for one and two doses and as a function of the specific circulating variant of the virus. We also assume that vaccinated individuals who get infected are less infectious by a factor accounting for the forward transmission reduction observed in vaccinated people [33]. Details of the mathematical definition and computational implementation of the model are reported in the Supplementary Information.

In Figure 2-A, we map the geographical location of the twenty LMIC covering all six WHO regions that were selected for modeling. Across the selected countries, we capture a wide range of vaccination coverage. By October 1, 2021, more than 50% of the populations in Sri Lanka, El Salvador, and Morocco had completed the initial COVID-19 vaccination course, compared to fewer than 3% in Ghana, Uganda, and Zambia. Countries such as the Philippines, Indonesia, Honduras, and Bolivia are in the middle with fractions between 19% and 24%. We calibrate the model in each country separately via an Approximate Bayesian Computation method [34] that uses as evidence the time series of recorded deaths in each country. This approach allows us to find the posterior distributions for a range of parameters such as the effective transmissibility of the different strains, seasonality, delay between deaths and their notification, under-reporting of deaths, and infection fatality rates (IFRs) [35] in the different countries as reported in the Supplementary Information. We first use the model to estimate for each country the number of deaths averted by the actual vaccination campaign, relative to a setting where no vaccines were available (results reported in the Supplementary Information). The model is run in the period 2020*/*12*/*01 − 2021*/*10*/*01, covering nearly one year of vaccine allocation and distribution, prior to the emergence of the Omicron variant. We refer the reader to the Supplementary Information for details about the calibration process and an estimate of the impact of the actual vaccination campaigns in averting deaths.

**Figure 2:**
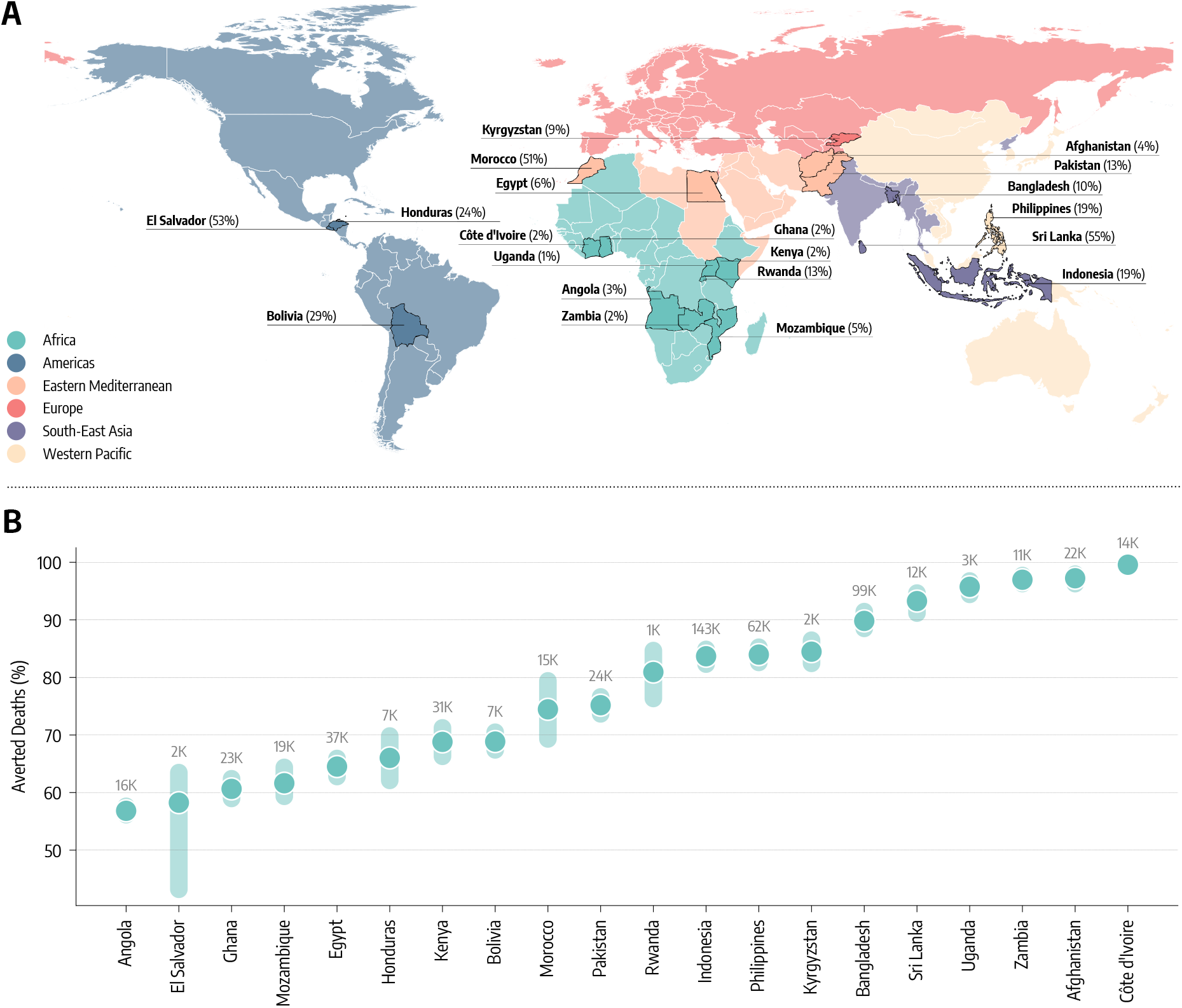
Counterfactual scenarios - Deaths averted if countries had US-equivalent vaccination rate. A) Countries modeled, their WHO region, and the percentage of fully vaccinated individuals there as of October 1, 2021. B) Deaths averted expressed as a percentage with respect to the actual vaccination rollout (median and interquartile range), assuming per capita vaccination rates equivalent to the United States. The median absolute number of deaths averted is reported above the inter-quartile range.

Our approach comes with the limitations of modeling studies. The details about the vaccination campaigns in some of the countries are limited. For example, the types of vaccines administered in some cases is missing. The model operates at a national level thus neglecting geographical heterogeneities that could reveal yet other layers of inequities, within each country. Although we consider variable IFRs across countries we do not explicitly account for comorbidities or limited healthcare access. Finally, the following counterfactual scenarios do not take into account the cost of the supply chain necessary to receive, store, distribute and administer doses.

## Counterfactual vaccination scenarios

We use the model to study counterfactual scenarios in which i) the per capita vaccination rate in the selected countries would have equaled that of the United States, and ii) vaccination rollout starts at the same time as in high-income countries, with no change in overall volume. In administering the extra available doses we assume a protocol that prioritizes the elderly population hence targeting a reduction of deaths rather than the reduction of the overall infection incidence [36–38]. For each counterfactual scenario, we estimate the impact of the changed vaccine rollout as the percent reduction in deaths (averted deaths) compared to the actual vaccine rollout.

In Figure 2-B, we report the estimated percentage of deaths averted and the median absolute numbers of deaths averted per country assuming the first counterfactual scenario of a US-equivalent vaccination rate. For half of the countries, the percentage of deaths averted exceeds 80%, with peaks close to 100% for Afghanistan, Côte D’Ivoire, and Zambia. In these countries, the absolute numbers of averted deaths are staggering, ranging from 143, 000 (IQR: [116, 000−183, 000]) in Indonesia to 3, 200 (IQR: [2, 100−5, 200]) in Uganda. For the other half of the countries, the percentages of deaths averted are in the 60% − 70% range, with absolute numbers ranging from 37, 800 (IQR: [24, 100 − 57, 100]) for Egypt to 2, 400 (IQR: [1, 600 − 3, 400]) in El Salvador.

The second counterfactual scenario investigates the impact of an earlier start of the vaccination campaign without changes in allocation volume. In other words, this scenario considers a more equal timing in the allocation of vaccine doses. We take as a reference point the start of the vaccination campaign in the US which took place on December 14, 2020. Among the countries under study, Indonesia administered the first doses in mid-January 2021, and Bangladesh, Egypt and Sri Lanka in late-January 2021. Eight countries started their vaccination in February: Bolivia and Pakistan in the beginning, El Salvador, Morocco, and Rwanda in the middle, Afghanistan, Honduras, and Philippines at the end of the month. Seven countries started their campaigns only in March: Angola, Cote d’Ivoire, Ghana, Mozambique and Uganda within the first ten days of the month while Kyrgyzstan at the end of it.

Finally, only one of the twenty countries under investigation (Zambia) started vaccinations in April (mid month). Hence, the delay with respect to the US starting date spans one to four months. Figure 3 shows how an earlier start would have been beneficial as it would have found a larger fraction of the population susceptible before the Delta wave. However, the magnitude of such effects is heterogeneous and overall smaller when compared to the previous counterfactual scenario with more vaccine doses. A key factor is the interplay between the amount of vaccine available and the relative shift of the starting time. Sri Lanka, El Salvador and Morocco, that achieved the highest coverage in the group and initiated their campaigns around two months later with respect to the US, would have averted more than 50% (73%, IQR: [69% − 76%] for Sri Lanka) of deaths. In the case of Angola and Zambia, both countries with a very low vaccination rate, a three/four month head start makes a difference (6, 100, IQR: [4, 200 − 8, 800] and 3, 400 [2, 300 − 4, 800] averted deaths respectively), but its effects are ultimately diminished by the relatively small number of administered vaccine doses. In fact, the percentage of averted deaths is very similar to what we find for Honduras that had a higher vaccination coverage but started only about two months later than the US. Similarly, the moderate percentage of averted deaths for Indonesia and Bangladesh, which however would have resulted in 36, 200 (IQR: [30, 000 − 46, 100]) and 22, 000 [15, 000 − 33, 500] fewer deaths, is due to the small difference between the actual and counterfactual start of the vaccination campaigns.

**Figure 3:**
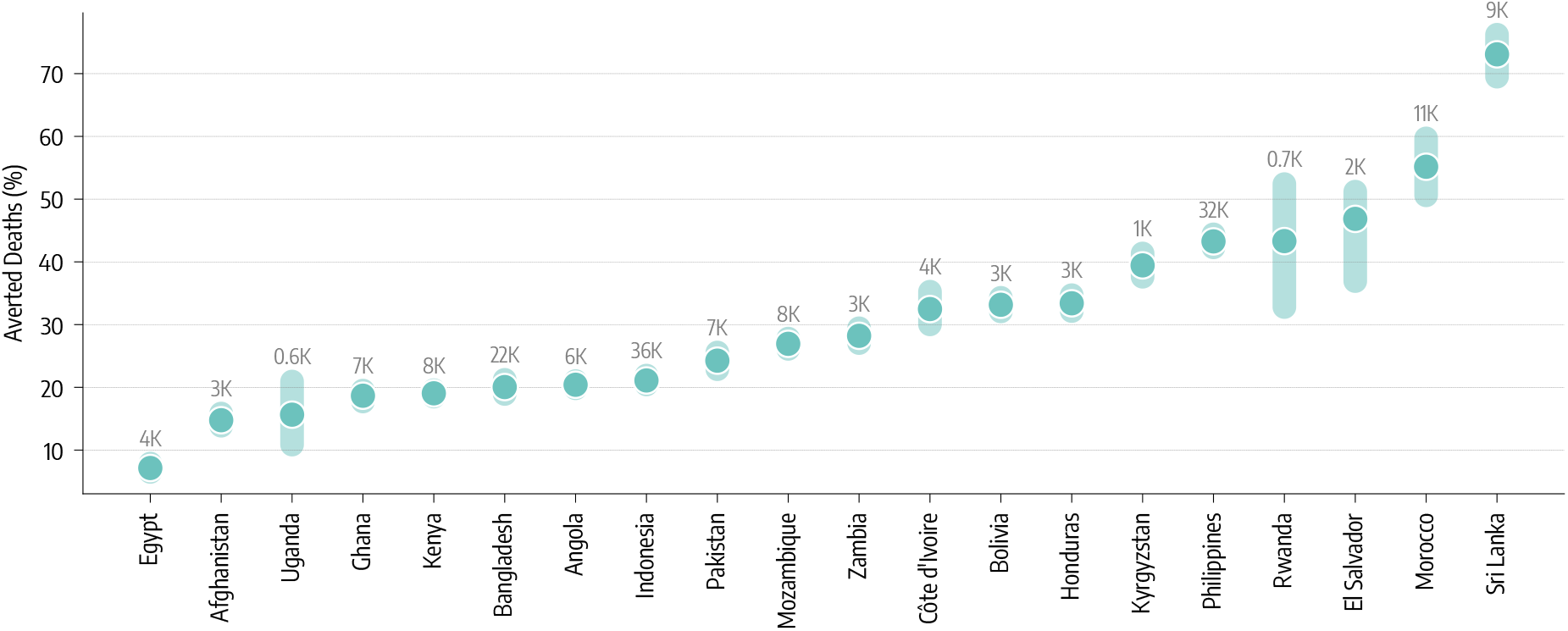
Counterfactual scenarios - Deaths averted if countries had US-equivalent vaccination start date. Deaths averted expressed as a percentage with respect to the actual vaccination rollout (median and interquartile range), assuming United States start date of December 14, 2020. The median absolute number of deaths averted is reported above the inter-quartile range.

## Estimate of NPIs required to offset vaccine inequity

In the counterfactual scenarios studied above, our modeling approach considers the same level of NPIs implemented in the factual scenario. For this reason we have investigated the extent of additional NPIs that would be needed to offset the inequitable allocation of vaccines. In this third counterfactual scenario we keep the doses administration as it unfolded in reality. Then, at week 51 of 2020, the start of the vaccination campaign in the US, we modify the NPIs to make them more restrictive. Since the impact of NPIs is modulated by their strictness and duration [15, 39–44], we explore a two-dimensional phase space in which NPIs are increased by *X*% with respect to the observed ones and the tightening of measures is maintained for *W* weeks. The percentage increase is defined as a corresponding decrease in contact rates (see Supplementary information for the modeling implementation). In Figure 4 we show the percentage increase in strictness of NPIs that, implemented for four months, would avert the equivalent numbers of deaths as achieved by a US-level vaccination rate. Notably, half of the countries would have needed NPIs increased by 40% or more during that period. The other half would need NPIs increased by around 30%. Ghana and Angola are the countries requiring the strongest NPIs to match the benefit of higher vaccination rates. As of October 1, 2021, they vaccinated only 2% and 3% of their population respectively. Although one might think that the key driver of the trend is the vaccination coverage, Pakistan, that in the same period vaccinated 13% of the people, is the country requiring the least NPIs to match the benefit of higher vaccination rates. Pakistan experienced a large epidemic wave in May 2021 and managed a rapid start of the vaccination campaign. Honduras instead started its campaign only a few weeks later but the initial vaccination rate was very slow (see the Supplementary Information for details). Indeed, the starting date and the rate of vaccination are equally important in defining the effectiveness of the vaccination campaigns. Furthermore, in this counterfactual scenario the increase of NPIs is implemented in different epidemiological contexts. The impact of NPIs has been critically linked to their timing in reference to the disease progression [15, 39–44]. For instance, Ghana experienced a big wave at the beginning of 2021, while Kyrgyzstan and Uganda, which are among the countries needing the least NPIs to match the effects of higher vaccination rate, experienced big waves only in the summer of 2021. For this reason in Figure 4 we also show a phase diagram for three specific countries detailing the averted deaths (color scale) as function of NPIs duration (x-axis) and increase (y-axis). Overall, we show that the longer additional NPIs are in place, the less strict they need to be to avert the same number of deaths. For example, Pakistan would have required an increase of about 30% of NPIs for two months to match the benefit brought by a higher vaccination rate. In contrast, in the case of the Philippines and Ghana we would need a 75% and 95% increase in NPIs in the same period.

**Figure 4:**
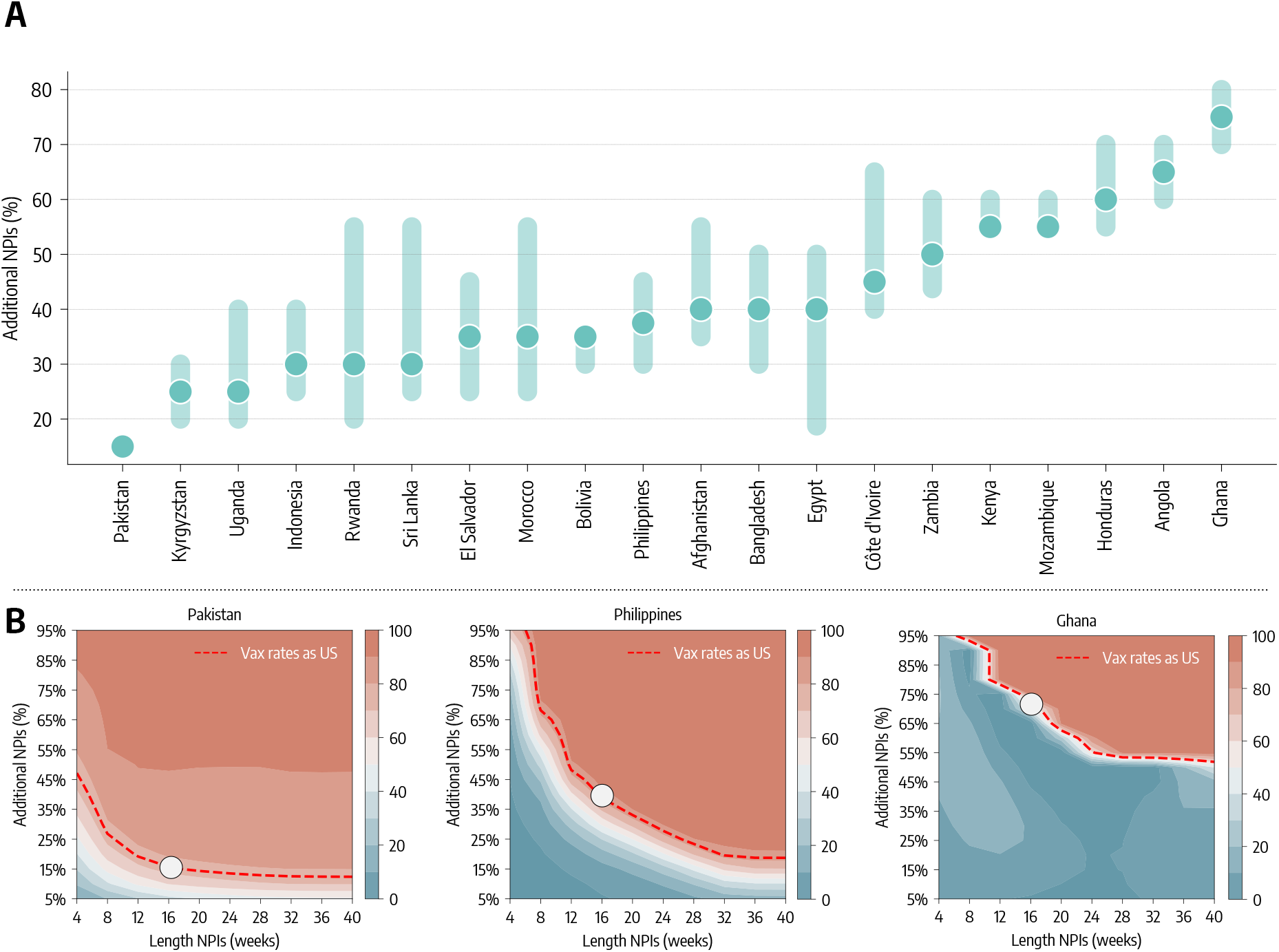
The role of NPIs. A) Additional NPIs, put in place for four months, needed to match the deaths averted that the vaccination rate of the US would have allowed. B) For three countries we show the contour plots of the percentage of deaths averted (median %) with stricter and/or longer NPIs, relative to the actual vaccination baseline. Percentage of deaths averted achieved by a US-equivalent vaccination rate is plotted as reference (red dashed line).

## Discussion

The overall picture emerging from our analysis shows that vaccine inequity, in both the number of doses available and the timeline of delivery, drastically reduced the impact of vaccination campaigns in the sample of LMIC studied. The emergence of new variants, featuring higher transmissibility and immune escape, suggests how vaccine inequity might become even more critical as we move forward in the pandemic [45]. Indeed, the original COVID-19 vaccines have been challenged by new variants (such as Omicron) when it comes to protection from infections and mild disease, but they still offer a very significant protection from severe outcomes. Progress in vaccine availability and administration in LMIC, also with respect to the administration of booster doses, would help considerably to mitigate the risks associated with highly transmissible new SARS-CoV-2 lineages. The presented approach and results can be extended to other countries and are potentially relevant in defining strategies aimed at minimizing the effect of inequities in vaccine allocation across countries.

## Data Availability

All data produced in the present study are available upon reasonable request to the authors

## Acknowledgements

MC and AV were partially supported by the Bill & Melinda Gates Foundation (award number INV006010). ND, MEH and IML acknowledge support from NIH-R01 AI139761. MEH, IML, and AV acknowledge support from the Ron Conway Family and the Emerson Collective. We acknowledge support from Google Cloud and Google Cloud Research Credits program. The content is solely the responsibility of the authors. All authors thank the High Performance Computing facilities at Greenwich University. N.G. acknowledges support from the DTA3/COFUND project funded by the European Union’s Horizon 2020 research innovation programme under the Marie Skłodowska Curie Actions grant agreement No 801604.

