## Supplementary Information for "Estimating the impact of COVID-19 vaccine allocation inequities: a modeling study"

### Supplementary Material for *Estimating the impact of COVID-19 vaccines allocation inequities: a modeling study*

#### Contents

**1 Data** **2**

**2 Epidemic model** **2**

**3 Model calibration** **4**

**4 Non-pharmaceutical interventions** **8**

**5 Modeling the introduction of a second SARS-CoV-2 strain** **9**

**6 Counterfactual scenarios** **9**

**7 Estimating the impact of the factual vaccination campaigns** **12**

### 1 Data

**Demographic and epidemic data.** Data about demographics comes from the United Nation World Population Prospects [1]. Epidemiological data are taken from the COVID-19 Data Repository by the Center for Systems Science and Engineering (CSSE) at Johns Hopkins University and from official sources [2].

**Vaccination data.** Data on global vaccine inequities used in the Introduction and in Fig. 1 of the main text are taken from the United Nations Development Programme via their Global Futures Platform [3]. Vaccination data used in the simulations are taken from Our World in Data [4]. The dataset provides the cumulative share of people partially ($p_t$) and fully ($f_t$) vaccinated against SARS-CoV-2 at time $t$. We turn these two quantities into daily number of administered doses. Consider the cumulative fraction of partially vaccinated individuals $p_t$. First, we fix possible null values using a linear interpolation. Second, we turn $p_t$ into the actual number of partially vaccinated $P_t$ by simply multiplying it by the total population $N$ of the country (i.e., $P_t = N \times p_t$). Finally, we take daily increases in the cumulative number of partially vaccinated individuals to get the number of daily first doses administered in the country. Through analogous calculations we get the number of daily second doses administered. In Fig. 1-A we show the percentage of individuals partially and fully vaccinated in the countries considered.

### 2 Epidemic model

We adopt a SLIR-like compartmental model (see Figure 2 for a schematic depiction). The susceptible individuals are placed in the compartment $S$. Getting in contact with the Infectious ($I$) they transition to the compartment of the Latent ($L$). Latent individuals are infected but become infectious only after $\epsilon^{-1}$ days when they eventually pass to the compartment $I$. After $\mu^{-1}$ days, infectious subjects finally transition to the compartment of the Recovered ($R$). By considering the COVID-19 characteristics we set $\epsilon^{-1} = 4days$ and $\mu^{-1} = 2.5days$. [5, 6]. We compute the number of deaths on daily recovered. In particular, the individuals that exit from the $I$ compartment, can either transition to the Recovered compartment ($R$) or the Dead compartment ($D$). The share of individuals transitioning to the $D$ compartment is regulated by the age-stratified Infection Fatality Rate (IFR) from Ref. [7]. To account for possible delays due to hospitalization and reporting between the transition $I \to R$ and actual death, we record the number of deaths computed on the recovered of day $t$ only after $\Delta$ days. In other words, $D$ individuals transition to the compartment $D^o$ (superscript $o$ stands for "observed") at a rate $1/\Delta$. Individuals are divided into 10 age groups ($0-9$, $10-19$, $20-24$, $25-29$, $30-39$, $40-49$, $50-59$, $60-69$, $70-79$, $80+$). The age-stratified rate of interaction are defined by the country specific contacts matrix $C$ from Ref. [8].

We also introduce a seasonal term to capture modulation of the force of infection regulated by changes in factors such as temperature and humidity [9, 10]. This means that in our simulation $R_t$ is multiplied by a rescaling factor $s_i(t)$ defined as $s_i(t) = \frac{1}{2}\left[\left(1 - \frac{\alpha_{min}}{\alpha_{max}}\right)\sin\left(\frac{2\pi}{365}(t - t_{max,i}) + \frac{\pi}{2}\right) + 1 + \frac{\alpha_{min}}{\alpha_{max}}\right]$, where $i$ refers to the hemisphere considered, and $t_{max,i}$ is the day associated to the maximum of the rescaling function. For the northern hemisphere it is set to January $15^{th}$ and to July $15^{th}$ for the southern hemisphere, while we consider no seasonal modulation in the tropical hemisphere. If a country extends across multiple zones, the seasonal factor is a weighted average of the different $s_i(t)$ according to the population living in the different hemispheres. We fix $\alpha_{max} = 1$ and consider $\alpha_{min}$ as a free parameter (see more details below).

In these settings, we model both vaccinations and the introduction of a second, more transmissible virus strain. In particular, individuals who received one dose of vaccine move to the compartments denoted with the superscript $V_1$. We assume that all individuals except for the infectious can receive the vaccine. Hence, susceptible, latent and recovered are vaccinated proportionally to their number. For $S^{V_1}$ individuals the force of infection is reduced by a factor $(1 - VE_{S1})$. If these individuals get infected, their IFR is also reduced by a factor $1 - VE_{M1}$. It follows that, in our simulations, the overall efficacy of a single dose of vaccine against death is $VE_1 = 1 - (1 - VE_{S1})(1 - VE_{M1})$. After receiving the second dose, individuals transition to the compartments with superscript $V_2$. Similarly, force of infection and IFR for them is reduced, respectively, by $(1 - VE_{S2})$ and $(1 - VE_{M2})$, implying an overall efficacy of $VE_2 = 1 - (1 - VE_{S2})(1 - VE_{M2})$. We also assume that vaccinated individuals that get infected are less infectious by a factor $(1 - VE_I)$ [11]. Since vaccine protection is not immediate, we introduce a delay of $\Delta_V$ days between administration (of both $1^{st}$ and $2^{nd}$ dose) and actual effect of the vaccine.

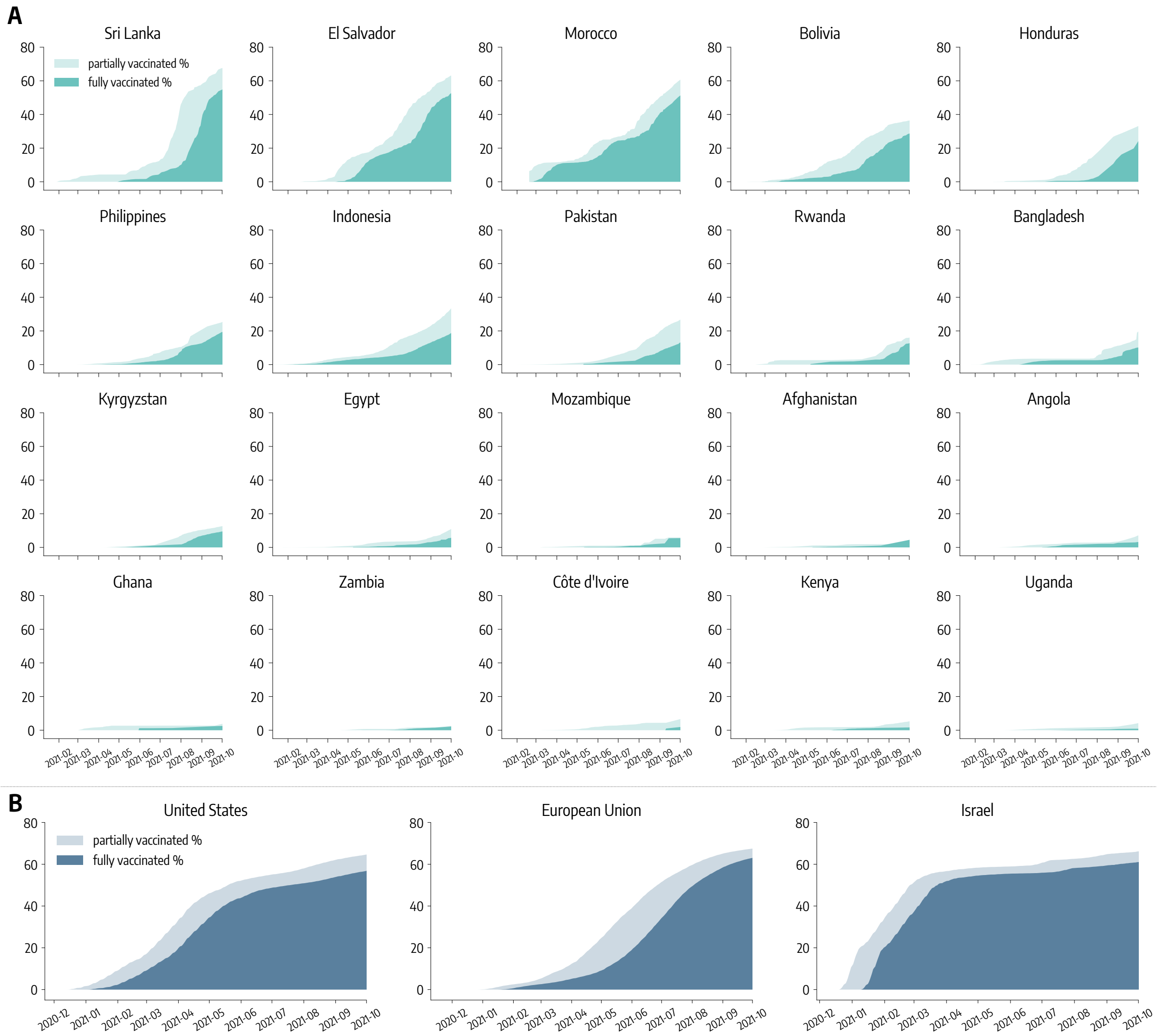

Figure 1: **Vaccinations.** A) Percentage of partially and fully vaccinated individuals in LMIC up to 2021/10/01. B) Percentage of partially and fully vaccinated individuals in high income settings up to 2021/10/01.

For example, an individual who received the $1^{st}$ dose on day $t$, will be protected with efficacy $VE_1$ only, on average, after $\Delta_V$ days. Hence, the transitions to compartments with superscript $V_1$ and $V_2$ take place at rate $\Delta_V^{-1}$ after first and second inoculation. We set $\Delta_V = 14days$. We do not have detailed information about the age of individuals receiving vaccines in all the countries considered. Therefore, we assume that the rollout proceeds prioritizing the elderly. We note how this is the strategy followed by the vast majority of governments worldwide [12–14]. This means that, in our model, vaccines are distributed in decreasing age order until all 50+ individuals are vaccinated, after vaccines are distributed homogeneously to the age groups $10-50$. We inform the model with the number of daily $1^{st}$ and $2^{nd}$ doses in different countries from Ref. [15]. In this work we set $VE_1 = 80\%$ ($VE_{S1} = 70\%$), $VE_2 = 90\%$ ($VE_{S2} = 80\%$), and $VE_I = 40\%$ [11].

We add specific $L$ and $I$ compartments to account for the introduction and emergence of a variant of concern. Considering the period under examination and the evidence from genomic surveillance in all countries under examination we consider the arrival and spread of the SARS-CoV-2 variant of concern Delta. Looking at genomic sequence data from Ref. [16–18] we get a proxy date for its introduction (more details provided below). We imagine that Delta is $\psi$ times more transmissible than the strain circulating previously and has a shorter latent period $\epsilon_{Delta}^{-1} = 3days$ [19]. We also assume that vaccines have a reduced efficacy against Delta VOC: $VE_1^{Delta} = 70\%$ ($VE_{S1}^{Delta} = 30\%$), $VE_2^{Delta} = 90\%$ ($VE_{S2}^{Delta} = 60\%$) [11].

The model is stochastic and transitions among compartments are simulated through chain binomial processes. More in detail, at time step $t$ the number of individuals in age group $k$ and compartment $X$ transiting to compartment $Y$ is sampled from $Pr^{Bin}(X_k(t), p_{X_k \to Y_k}(t))$, where $p_{X_k \to Y_k}(t)$ is the transition probability. As an illustrative example consider the number of new infected individuals from the $S$ compartment. The rate for this transition is called *force of infection* (generally referred as $\lambda$) and may depend on several factors, from transmissibility of the pathogen to contact rates and seasonality. In our case, the force of infection for age group $k$ at time $t$ is defined as:

$$\lambda_k(t) = \beta \times s(t) \times r(t) \times \left\{ \sum_{k'} \frac{C_{kk'}}{N_{k'}} \left[ I_{k'} + (1 - VE_I)(I_{k'}^{V1} + I_{k'}^{V2}) \right] \right\} \tag{1}$$

Where $\beta$ is the transmission rate, $s(t)$ is the seasonality factor, $r(t)$ captures contacts reduction due to NPIs (more details below), and the term in brackets is the probability of contacting an infectious individual in age group $k'$ given the contact rates and the number of individuals per group. Notice how infectious individuals that received a vaccine (either one or two dose) have their infectiousness reduced by the $(1 - VE_I)$ factor. The probability of the infection transition is simply the force of infection multiplied by the length of the simulation step $\Delta t$ (i.e., $p_{S_k \to L_k}(t) = \lambda_k(t)\Delta t$). Here, the unit of time of the simulation is the day, therefore $\Delta t = 1$. The number of $S_k$ individuals getting infected at time $t$ is then extracted from $Pr^{Bin}(S_k(t), \lambda_k(t))$. Given this, we can easily get the infection probability also for susceptible individuals that received one or two vaccine doses, namely $S^{V_1}$ and $S^{V_2}$. Indeed, since vaccines offer a protection $VE_S$ from infection, we have that $p_{S_k^{V_1} \to L_k^{V1}}(t) = (1 - VES_1) \times \lambda_k(t)$ and $p_{S_k^{V_2} \to L_k^{V2}}(t) = (1 - VES_2) \times \lambda_k(t)$.

### 3 Model calibration

The free parameters of the model are calibrated through a rejection algorithm based on Approximate Bayesian Computation [20, 21]. We define a prior distribution $P(\theta)$ on the free parameters $\theta$. At each step of the iterative algorithm, we sample a set of parameters $\hat{\theta}$ from $P(\theta)$ and an instance of the model is generated using $\hat{\theta}$. Then, an output quantity of the model $E'$ is compared to the corresponding real quantity $E$ using an error metric $S(E, E')$: if $S(E, E')$ is smaller than a tolerance $\delta$, $\hat{\theta}$ is accepted otherwise is rejected. Repeating this procedure iteratively we obtain an approximation of the posterior distribution of the parameters $\theta$. In this work we consider weekly deaths as output quantity and the weighted mean absolute percentage error (wMAPE) as distance metric. The free parameters and the relative priors are:

- the transmission rate $\beta$; we explore uniformly values such that the $R_t$ on the first simulation date is between 0.6 and 2.0;
- the delay in deaths $\Delta \sim U(10, 35)$ [22];

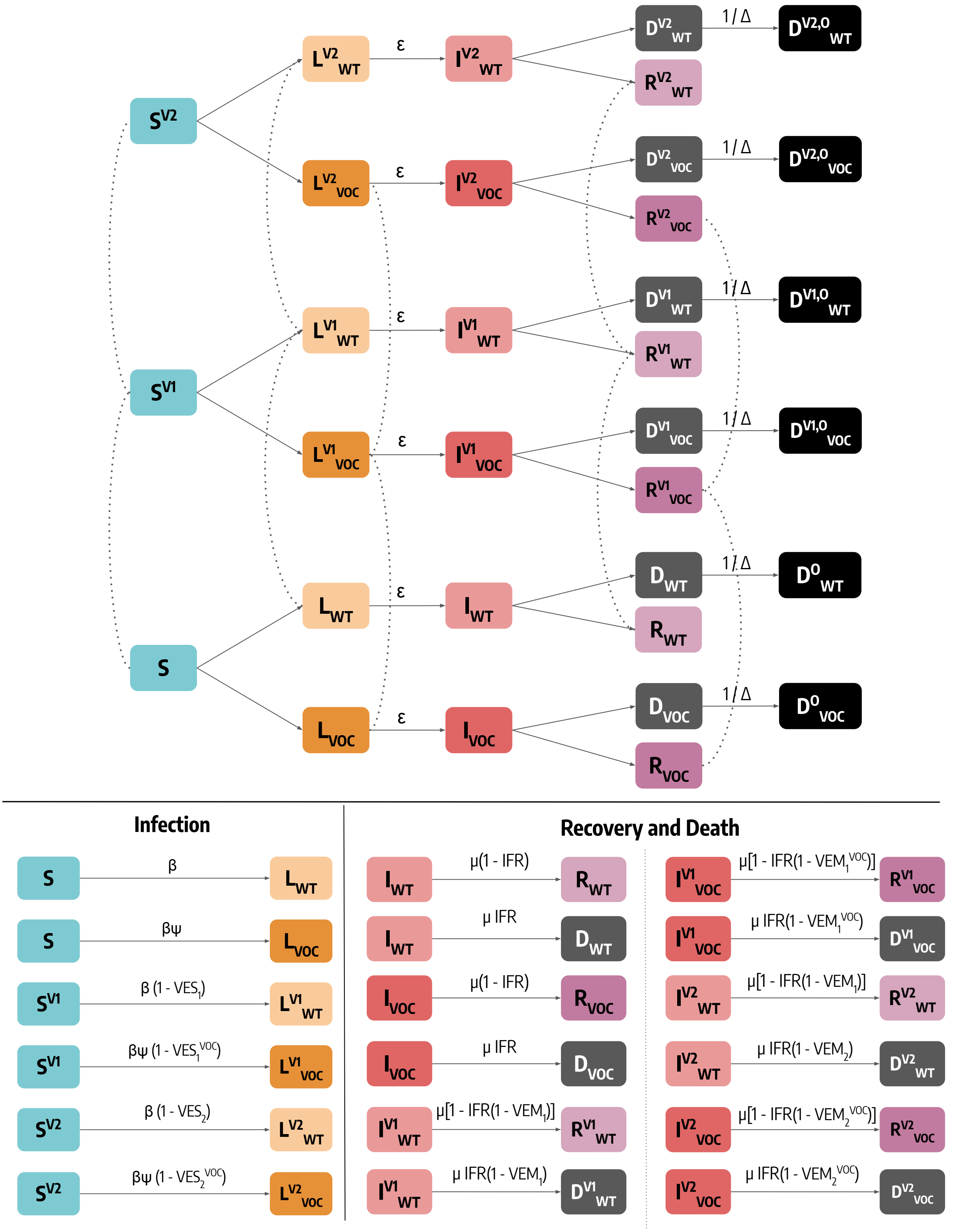

Figure 2: **Schematic representation of the epidemic model.** For simplicity, we represent the model for a single age group. Dashed lines indicate data-driven transitions linked to vaccination status, solid lines indicate that simulated transitions. In the bottom of the figure we report the rate of transitions related to both infection and and recovery/death.

- the seasonality parameter $\alpha_{min} \sim U(0.5, 1.0)$ (0.5 indicates strong seasonality while 1.0 absence of seasonality);
- the initial number of infected individuals; we explore uniformly values between 1 and 25 times the number of cases notified in the 7 days prior the beginning of the simulation ($Inf^{mult}_{start}$). We divide these individuals in the infected compartments ($L$, $I$) proportionally to the time spent there by individuals ($\epsilon^{-1}$ for $L$ and $\mu^{-1}$ for $I$);
- the initial number of recovered; we explore uniformly values between 1 and 25 times the total number of reported cases up to the start of the simulation ($Rec^{mult}_{start}$);
- the relative transmissibility advantage of the Delta VOC $\psi \sim U(1.0, 2.5)$
- the date of the introduction of the Delta VOC. We consider values between 1 month before and after the date when Delta was responsible for at least 5% of sequenced samples according to the data from Ref. [16] (more details below);
- the IFR multiplier $\sim U(0.5, 2.0)$; this number multiplies the IFR from Ref. [7];
- the percentage of deaths reported $\sim U(5\%, 100\%)$;

The model is calibrated separately for different countries during the period 2020/12/01 – 2021/10/01. Here, instead of defining a tolerance, For a single country we run $1M$ simulations, and we keep the 1000 parameter samples that achieve the lowest wMAPEs. These sets represent the posterior distributions and are used to run the calibrated models. In Tab. 1, 2, 3, 4, 5 we report the posterior distributions (median and interquartile range) of the free parameters.

| | **Sri Lanka** | **El Salvador** | **Morocco** | **Bolivia** |
|---|---|---|---|---|
| $R_t^{start}$ | 0.89 [0.86, 0.91] | 1.05 [1.0, 1.11] | 1.21 [1.12, 1.31] | 1.19 [1.13, 1.28] |
| $\Delta$ | 14 [12, 17] | 26 [12, 32] | 12 [11, 15] | 20 [19, 22] |
| $\alpha_{min}$ | 0.75 [0.61, 0.89] | 0.76 [0.62, 0.86] | 0.82 [0.71, 0.93] | 0.75 [0.63, 0.88] |
| $\psi$ | 2.43 [2.34, 2.47] | 1.58 [1.4, 1.71] | 1.81 [1.65, 2.12] | 1.52 [1.23, 1.93] |
| $Inf^{mult}_{start}$ | 2.93 [1.84, 4.81] | 24.04 [23.33, 24.58] | 6.03 [3.94, 9.12] | 14.4 [13.11, 15.75] |
| $Rec^{mult}_{start}$ | 10.04 [5.47, 16.89] | 19.79 [15.12, 22.58] | 14.08 [8.17, 19.85] | 8.59 [4.8, 13.83] |
| *IFR Multiplier* | 1.1 [0.78, 1.49] | 1.18 [0.8, 1.62] | 1.05 [0.73, 1.43] | 1.81 [1.69, 1.91] |
| *% deaths reported* | 51.6 [35.33, 71.19] | 40.74 [27.06, 64.1] | 33.25 [24.13, 48.22] | 89.73 [83.82, 95.12] |
| *Date intro. VOC* | 05-02 [05-01, 05-04] | 06-06 [05-28, 06-12] | 05-02 [04-28, 05-08] | 08-23 [08-10, 09-02] |

Table 1: **Posterior distributions of free parameters obtained via ABC calibration (Sri Lanka, El Salvador, Morocco, Bolivia)**. We show median and interquartile range of the different parameters. Dates are represented with a $mm - dd$ format and refer all to the year 2021.

| | **Honduras** | **Philippines** | **Indonesia** | **Pakistan** |
|---|---|---|---|---|
| $R_t^{start}$ | 1.44 [1.35, 1.53] | 1.35 [1.31, 1.39] | 1.23 [1.21, 1.25] | 0.97 [0.96, 0.99] |
| $\Delta$ | 32 [29, 33] | 33 [25, 34] | 29 [27, 32] | 24 [21, 26] |
| $\alpha_{min}$ | 0.74 [0.63, 0.87] | 0.75 [0.63, 0.87] | 0.72 [0.62, 0.86] | 0.88 [0.86, 0.9] |
| $\psi$ | 1.74 [1.65, 1.8] | 1.53 [1.24, 1.77] | 1.51 [1.38, 1.67] | 1.12 [1.07, 1.16] |
| $Inf_{start}^{mult}$ | 9.25 [8.29, 10.87] | 5.82 [4.17, 7.31] | 14.53 [11.0, 17.73] | 16.12 [12.02, 20.69] |
| $Rec_{start}^{mult}$ | 11.5 [6.69, 17.49] | 13.81 [7.03, 18.7] | 12.74 [5.9, 17.89] | 13.07 [7.68, 19.43] |
| *IFR Multiplier* | 1.2 [0.87, 1.54] | 1.0 [0.71, 1.41] | 1.35 [1.05, 1.7] | 1.25 [0.91, 1.62] |
| *% deaths reported* | 54.41 [41.19, 74.39] | 31.54 [22.57, 45.36] | 69.12 [52.71, 84.26] | 55.17 [39.26, 75.44] |
| *Date intro. VOC* | 06-09 [06-03, 06-12] | 06-29 [06-20, 07-04] | 05-11 [04-27, 05-18] | 05-26 [05-08, 06-05] |

Table 2: **Posterior distributions of free parameters obtained via ABC calibration (Honduras, Philippines, Indonesia, Pakistan)**. We show median and interquartile range of the different parameters. Dates are represented with a $mm - dd$ format and refer all to the year 2021.

| | **Rwanda** | **Bangladesh** | **Kyrgyzstan** | **Egypt** |
|---|---|---|---|---|
| $R_t^{start}$ | 1.35 [1.33, 1.36] | 1.11 [1.08, 1.14] | 1.4 [1.31, 1.51] | 1.17 [1.12, 1.2] |
| $\Delta$ | 13 [12, 15] | 19 [17, 21] | 28 [25, 32] | 21 [16, 25] |
| $\alpha_{min}$ | 0.75 [0.63, 0.88] | 0.92 [0.84, 0.96] | 0.91 [0.83, 0.95] | 0.8 [0.74, 0.85] |
| $\psi$ | 1.07 [1.03, 1.13] | 1.54 [1.41, 1.69] | 1.27 [1.15, 1.4] | 1.16 [1.08, 1.24] |
| $Inf_{start}^{mult}$ | 18.79 [14.32, 21.82] | 7.37 [3.68, 12.45] | 2.48 [1.81, 3.23] | 21.59 [18.35, 23.37] |
| $Rec_{start}^{mult}$ | 12.89 [7.68, 17.9] | 12.9 [6.72, 18.31] | 13.19 [7.39, 19.07] | 12.35 [6.76, 18.97] |
| *IFR Multiplier* | 1.3 [0.92, 1.62] | 1.05 [0.71, 1.46] | 1.25 [0.9, 1.63] | 1.16 [0.77, 1.65] |
| *% deaths reported* | 56.07 [40.37, 74.8] | 16.92 [11.28, 25.17] | 42.39 [30.42, 59.53] | 14.95 [10.04, 23.69] |
| *Date intro. VOC* | 05-02 [04-19, 05-25] | 04-11 [03-28, 04-23] | 04-03 [03-19, 04-13] | 05-17 [04-28, 06-02] |

Table 3: **Posterior distributions of free parameters obtained via ABC calibration (Rwanda, Bangladesh, Kyrgyzstan, Egypt)**. We show median and interquartile range of the different parameters. Dates are represented with a $mm - dd$ format and refer all to the year 2021.

|  | **Mozambique** | **Afghanistan** | **Angola** | **Ghana** |
|---|---|---|---|---|
| $R_t^{start}$ | 1.59 [1.56, 1.63] | 1.34 [1.29, 1.38] | 1.56 [1.54, 1.58] | 1.58 [1.55, 1.61] |
| $\Delta$ | 28 [26, 31] | 12 [11, 15] | 31 [29, 33] | 15 [12, 18] |
| $\alpha_{min}$ | 0.72 [0.61, 0.85] | 0.93 [0.83, 0.97] | 0.74 [0.62, 0.87] | 0.75 [0.62, 0.87] |
| $\psi$ | 1.39 [1.3, 1.47] | 1.48 [1.3, 1.62] | 2.18 [2.03, 2.32] | 1.98 [1.87, 2.1] |
| $Inf_{start}^{mult}$ | 20.71 [17.64, 22.87] | 3.41 [1.67, 8.06] | 19.69 [15.99, 22.6] | 19.76 [15.97, 22.5] |
| $Rec_{start}^{mult}$ | 12.95 [7.34, 19.1] | 12.67 [6.76, 18.84] | 12.19 [6.29, 18.62] | 13.82 [7.3, 19.38] |
| *IFR Multiplier* | 1.0 [0.72, 1.43] | 0.97 [0.7, 1.39] | 1.04 [0.71, 1.45] | 0.91 [0.67, 1.26] |
| *% deaths reported* | 5.58 [3.88, 7.7] | 19.9 [13.59, 27.68] | 2.32 [1.66, 3.31] | 1.83 [1.35, 2.51] |
| *Date intro. VOC* | 04-22 [04-08, 05-04] | 04-12 [04-07, 04-28] | 05-31 [05-26, 06-03] | 05-26 [05-18, 06-02] |

Table 4: **Posterior distributions of free parameters obtained via ABC calibration (Mozambique, Afghanistan, Angola, Ghana)**. We show median and interquartile range of the different parameters. Dates are represented with a $mm - dd$ format and refer all to the year 2021.

|  | **Zambia** | **Côte d'Ivoire** | **Kenya** | **Uganda** |
|---|---|---|---|---|
| $R_t^{start}$ | 1.25 [1.24, 1.27] | 1.08 [1.06, 1.09] | 1.38 [1.37, 1.41] | 1.41 [1.39, 1.44] |
| $\Delta$ | 12 [11, 14] | 27 [19, 31] | 30 [27, 32] | 30 [28, 31] |
| $\alpha_{min}$ | 0.73 [0.62, 0.86] | 0.77 [0.64, 0.88] | 0.74 [0.62, 0.89] | 0.76 [0.64, 0.86] |
| $\psi$ | 1.13 [1.08, 1.2] | 1.09 [1.05, 1.15] | 1.41 [1.36, 1.47] | 1.11 [1.06, 1.18] |
| $Inf_{start}^{mult}$ | 16.27 [10.17, 21.0] | 16.07 [9.63, 20.61] | 20.5 [17.8, 22.96] | 1.62 [1.27, 2.14] |
| $Rec_{start}^{mult}$ | 13.13 [7.4, 19.23] | 13.19 [7.32, 19.05] | 13.02 [7.93, 19.22] | 13.84 [7.91, 19.86] |
| *IFR Multiplier* | 1.05 [0.71, 1.46] | 1.0 [0.73, 1.39] | 0.99 [0.72, 1.38] | 1.04 [0.77, 1.43] |
| *% deaths reported* | 21.87 [15.94, 31.72] | 2.11 [1.57, 2.96] | 6.58 [4.78, 9.31] | 42.36 [26.11, 63.94] |
| *Date intro. VOC* | 04-08 [03-22, 04-21] | 05-31 [05-13, 06-14] | 05-01 [04-23, 05-07] | 03-17 [03-01, 03-31] |

Table 5: **Posterior distributions of free parameters obtained via ABC calibration (Zambia, Côte d'Ivoire, Kenya, Uganda)**. We show median and interquartile range of the different parameters. Dates are represented with a $mm - dd$ format and refer all to the year 2021.

### 4 Non-pharmaceutical interventions

We model the effects of non-pharmaceutical interventions (NPIs) on contacts using the COVID-19 Community Mobility Report By Google [23]. The dataset provides, for various countries and spatial resolutions, a percentage change in mobility $r(t)$ on day $t$. We convert this quantity into a contacts reduction parameters $c(t)$ following the relation: $c(t) = (1 + r(t)/100)^2$. Indeed, the number of contacts scale with the square of the number of individuals. For example, a percentage reduction of $-20\%$ translates into a contacts reduction factor of 0.64. In the simulations the contacts matrix $C$ is multiplied by this reduction parameter $c(t)$ to account for the modulation in contacts induced by NPIs. The dataset provides mobility changes with respect to specific locations. In this work, we compute $r(t)$ by using the average of the fields `workplaces percent change from baseline`, `retail and recreation percent change from baseline` and `transit stations percent change from baseline`.

### 5 Modeling the introduction of a second SARS-CoV-2 strain

We model the introduction of a second SARS-CoV-2 strain considering genomic sequencing data from Co-Variants [16]. The data provides the fraction of processed samples by virus variant in different countries. As clear from the plots, in the period and countries under examination we observed the introduction and rapid growth of the Delta variant of concern. To reduce the impact of noise on seeding date estimates, for each country, we fit a logistic curve of the type $\hat{s}(t) = 1/(1 + e^{-\gamma(t - t_{1/2})})$ to the real fractions of Delta variant samples $s(t)$. The fit is performed via least square using the python library *scipy* [24]. In Fig. 3 we show the actual Delta prevalence (i.e., fraction of samples that fall into the Delta group) to the fitted prevalence. We also show the total number of samples processed in each week. In the case of countries for which genomic data is not available, we perform the logistic fit on all the samples from neighbouring countries for which data is available.

After the fit, we pick the first date on which the fitted Delta prevalence is greater or equal to 5% (i.e., $t \mid \hat{s}(t) \geq 0.05$). When running the simulations, on that date we calculate the 5% of the daily simulated infected individuals and we use them to initialize the compartment of the infected with the Delta variant. We choose a 5% threshold to avoid a prevalence that is too low and therefore more affected by noise. At the same time, we did not choose a higher prevalence threshold to avoid imposing a strong discontinuity on the $R_t$ on the simulations.

### 6 Counterfactual scenarios

#### 6.1 Vaccination rates of high income settings

We propose scenarios in which the LMIC considered manage the same vaccines availability of three high income contexts: United States (shown in the main text), European Union, and Israel (shown here). To do so, we run simulations in which, instead of the factual vaccination data of the LMIC considered, we use the daily number of first and second doses administered in the three high income settings. To account for different population sizes among geographical regions, we rescale the number of doses available in the counterfactual. For example, consider the case when we apply to Mozambique the vaccination rates of Israel. If Israel administered $X_t^{Israel}$ doses on day $t$, in the counterfactual scenario we administer $X_t'^{Mozambique} = X_t^{Israel} \frac{N_{Mozambique}}{N_{Israel}}$. In Fig. 1-B we show the percentage of partially and fully vaccinated in the three high income settings. Vaccinations started on 2020/12/14 in US, on 2020/12/27 in most of European countries, and on 2020/12/19 in Israel. As of 2021/10/01, the European Union shows the highest percentage of fully vaccinated (63%), followed by Israel (61%), and US (57%). Nonetheless, we acknowledge differences among the three vaccination rollout especially at the beginning. Indeed, we see that in Israel and, to a lower extent also in the US, vaccinations were much faster after the start respect to the European Union. For example, the percentage of fully vaccinated on the 2021/03/01 was: 2.6%, 9.2%, and 37.3% in, respectively, EU, US, and Israel.

In Fig. 4, we show the percentage of deaths that are averted applying to LMIC the vaccination rates of US, EU, and Israel with respect to simulations with factual doses. The overall picture presented in the main text for US vaccination rates holds also with EU and Israel rates. Indeed, in both cases we observe that additional doses of vaccine bring a huge benefit in terms of reduction of fatalities. When considering dose availability of Israel, averted deaths in LMIC span between 60% to nearly 100%, while these figures lie between 40% and 90% when EU rates are considered. We also note that the ordering of the countries is the same when considering the three different rates. Across the LMIC considered, the greatest decrease in deaths is achieved by Israel rates, while EU rates are the least effective at reducing the number of deaths. A possible explanation is that, as noted previously, while EU reached a higher vaccine coverage as of 2021/10/01, the vaccine rollout in Israel was much faster in the early months of 2021, allowing to provide significant level of protection at the population level quicker. The results obtained with US rates lie in the middle between those obtained with Israel and EU rates. In light of the previous explanation, this makes sense: the early rollout in US was quicker than EU but slower than Israel.

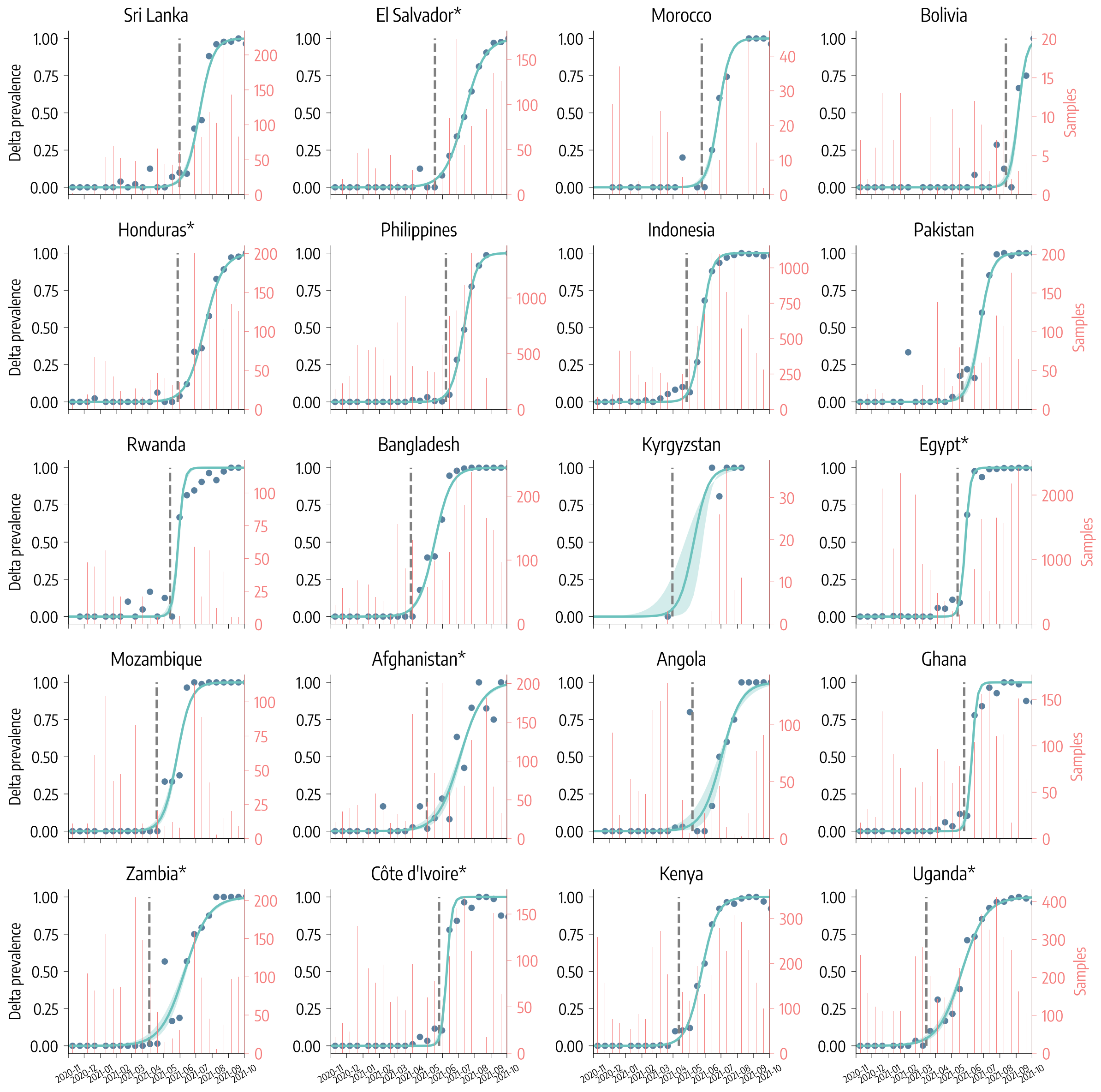

Figure 3: **Introduction of Delta variant.** We represent the actual fraction of Delta samples and the fitted Delta prevalence. We also display the total number of samples processed per week. For countries denoted by * data are not available, therefore in those cases the fit is performed on all samples from neighbouring countries for which data are available.

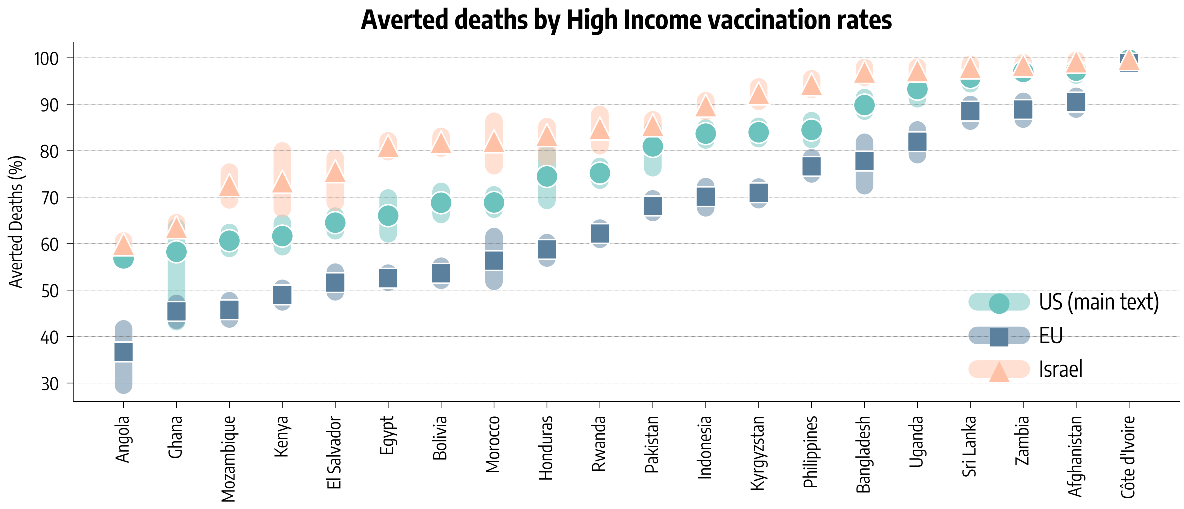

Figure 4: **Averted Deaths with High Income regions vaccination rates.** Averted deaths (median and inter-quartile range) expressed as a percentage with respect to the factual vaccination baseline using the vaccination rates of the United States, European Union, and Israel.

#### 6.2 Earlier start of factual vaccinations

As a second counterfactual analysis, we anticipate the factual vaccination campaign in the LMIC in order to match the start of vaccine rollout in high income settings. As new starting date we choose the 2020/12/14, when COVID-19 vaccinations started in United States. If the shift of vaccination data causes missing data at the end of the time series of LMIC, we fill it considering the average number of doses administered during the last 7 days. It is important to stress how this counterfactual does not increases the number of doses nor the rate of vaccination. It is a solid shift of the starting date to an earlier point.

#### 6.3 NPIs increase

We consider simulations in which we modify the factual NPIs quantified with the COVID-19 Community Mobility Report. More in detail, given the contacts reduction factor of week $t$ $c(t)$, in the new simulations with $X\%$ additional NPIs the new factor will be $c'(t) = c(t)(1 - X/100)$. The increase of NPIs is modeled as a contacts reduction factors after week 51 of 2020, a proxy date for the start of vaccinations in the United States. Indeed, our goal is to estimate the additional amount of NPIs needed to match the number of deaths averted when applying vaccination rates of a high income setting such as US. Finally, the increase of NPIs is sustained for a limited number of weeks. We explore multiple scenarios with NPIs that are increased in the range of 5% to 95% and that are sustained for a number of weeks between 4 and 40. We run additional simulations in which we modify the NPIs as described and we compute the fraction of averted deaths with respect to simulations with factual NPIs and vaccine rollout.

### 7 Estimating the impact of the factual vaccination campaigns

In Figure 1-A we have shown the evolution of the percentage of partially and fully vaccinated in the twenty LMIC countries up to 2021/10/01. As clear from the graphs, there is a high level of heterogeneity. We go from fractions of fully vaccinated above 50% in Sri Lanka and Morocco, to values below 10% in Kyrgyzstan, Mozambique, Egypt, and even below 1% in Uganda. Honduras, Bolivia, Indonesia are around the middle between these two groups with a fraction of fully vaccinated between 20% and 30%. When interpreting the numbers it is important to recognize the differences in terms of populations. Indonesia and Pakistan are the largest with 273M and 220M people respectively. El Salvador and Kyrgyzstan are the smallest with around 6M residents each. Hence, the differences in terms of the absolute number of vaccinated individuals and doses administered span several orders of magnitudes among these countries.

In Figure 5, we show the real data of confirmed deaths (dark blue dots). In most countries, the latest epidemic wave, caused by the Delta variant, was, unfortunately, the most deadly. Clearly in contrast with observations across high income countries where, despite the increased transmissibility and severity, the Delta wave was strongly limited by high vaccination rates with respect to the previous [25, 26]. This observation is a first clear hint about the impact that vaccines could have had in these settings. In the plots, we also report the median and confidence intervals of our fits (light green lines and shaded areas). Across the board, the model can capture the evolution of the pandemic with accuracy. It is however important to highlight the few misses. In the case of Sri Lanka the model produces a peak that is slightly delayed with respect to observations. In the case of Zambia and Côte d'Ivoire, the model fails to capture the surge in deaths in early 2021. Each plot reports also the model's prediction of what would have happened in total absence of vaccines (red dashed lines and shaded areas). In particular, we run the model keeping all the same fitted parameters, NPIs, but remove all doses administered. In doing so, we provide estimates of the impact of the factual vaccines in each country. Again, we find large heterogeneity induced by the radically different vaccination coverage. Countries that managed to vaccinate more, such as Sri Lanka, El Salvador, and Morocco show the largest differences between the real evolution of confirmed deaths and those in the hypothetical scenario without vaccines (i.e., baseline). Conversely, in countries such as Kenya, and Uganda, that have a minimal vaccination coverage, the differences are very small. Figure 6 confirms this picture but provides a more clear estimation of the impact of vaccines. We plot the deaths averted by the factual vaccination campaigns (median) versus the fraction of individuals fully vaccinated as of 2021/10/01 in the LMIC considered. Here, averted deaths are computed with respect to the baseline simulations without vaccines administered. We observe a high correlation between averted deaths and vaccination coverage (Pearson coefficient $r = 0.93$, $p_{value} < 0.001$). In Sri Lanka, Morocco and El Salvador the vaccine rollout averted about 70% of the deaths with respect to the baseline. In Bolivia, Indonesia, Honduras, and Philippines the numbers are lower but still significant. Finally, we observe the group of countries where the doses administered are very limited ($< 10\%$) but their impact is still positive and not negligible. While interpreting the results and comparing countries it is important to stress how the model is fitted separately to each nation. Hence, some values of the free parameters such as the effective transmissibility of the strains circulating might be estimated as slightly different even though they refer to the same variants. For example, the posterior distribution for the relative transmissibility advantage of the Delta variant with respect to Alpha peaks at 1.8 in Morocco while at 2.4 in Sri Lanka. These are effective parameters selected based on the available data. As such, they factor in many behavioral factors that are not explicitly modeled. Examples are the relations among mobility reduction, contact rates modifications, and infections. These might differ in different contexts/environments and affect the scenarios modelled here.

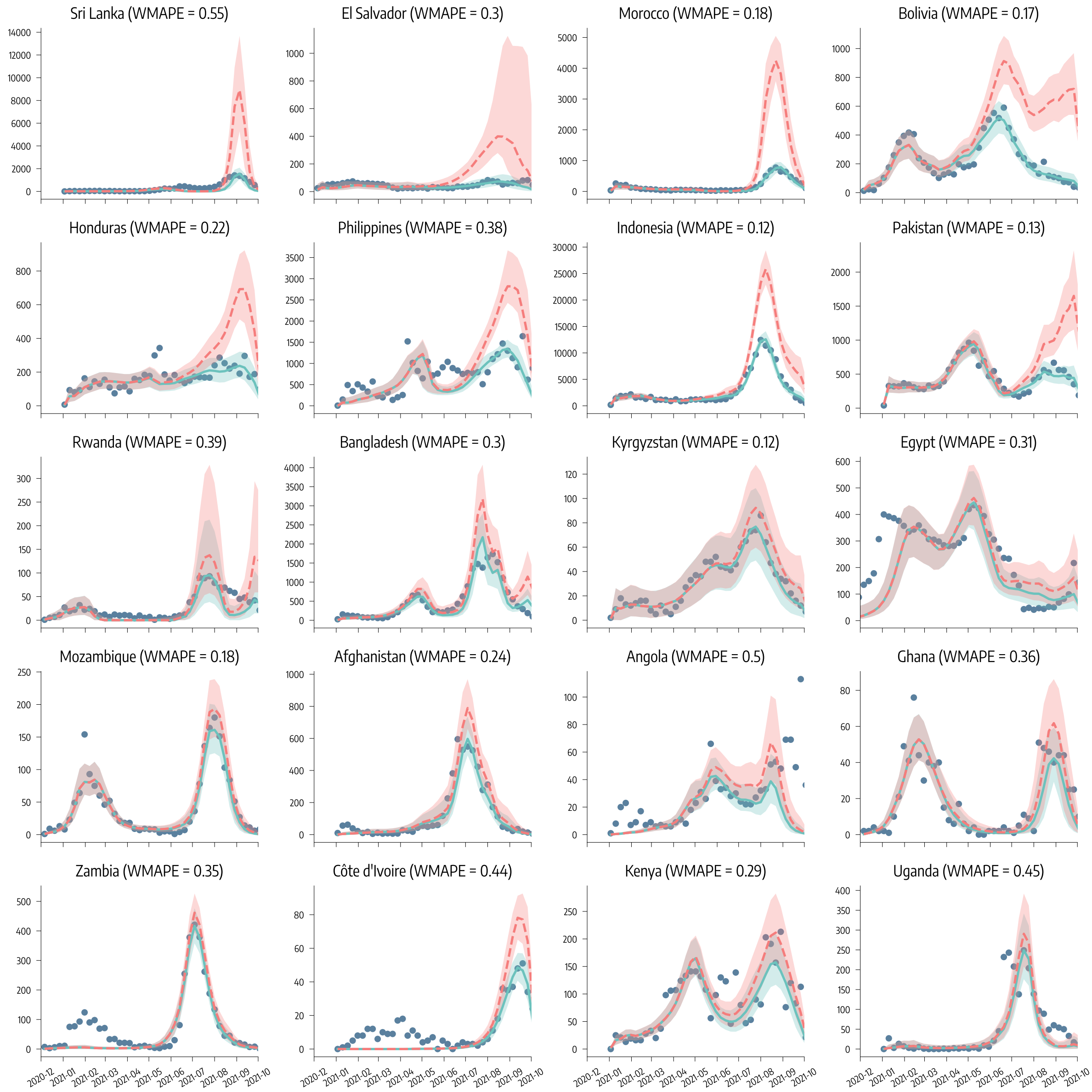

Figure 5: **Calibration results and impact of factual vaccinations in LMIC**. We show the actual number of weekly deaths (blue dots) and the simulated weekly deaths via the calibrated model (median and 90% CI). We also show the estimated number of weekly deaths in absence of vaccines (red dashed line).

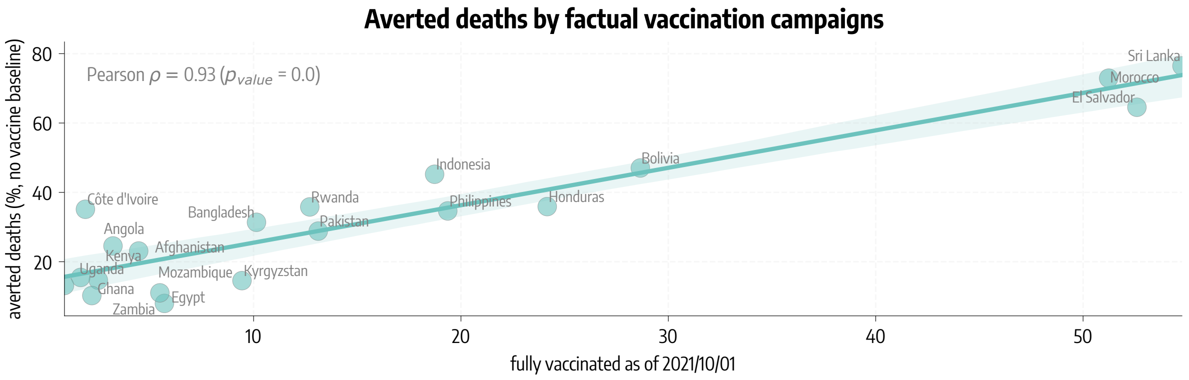

Figure 6: **Deaths averted by factual vaccination campaigns in LMIC**. We show the estimated percentage of averted deaths (median) with respect to baseline simulations without vaccines versus the percentage of individuals fully vaccinated as of 2021/10/01 in the LMIC considered.

[7] Robert Verity, Lucy Okell, Ilaria Dorigatti, Peter Winskill, Charles Whittaker, Natsuko Imai, Gina Cuomo-Dannenburg, Hayley Thompson, Patrick Walker, Han Fu, Amy Dighe, Jamie Griffin, Marc Baguelin, Sangeeta Bhatia, Adhiratha Boonyasiri, Anne Cori, Zulma M. Cucunubá, Rich FitzJohn, Katy Gaythorpe, and Neil Ferguson. Estimates of the severity of coronavirus disease 2019: a model-based analysis. *The Lancet Infectious Diseases*, 20, 03 2020.

[8] Dina Mistry, Maria Litvinova, Ana Pastore y Piontti, Matteo Chinazzi, Laura Fumanelli, Marcelo F C Gomes, Syed A Haque, Quan-Hui Liu, Kunpeng Mu, Xinyue Xiong, M Elizabeth Halloran, Ira M Longini, Stefano Merler, Marco Ajelli, and Alessandro Vespignani. Inferring high-resolution human mixing patterns for disease modeling. *Nature Communications*, 12(1):323, 2021.

[9] Duygu Balcan, Bruno Gonçalves, Hao Hu, José J. Ramasco, Vittoria Colizza, and Alessandro Vespignani. Modeling the spatial spread of infectious diseases: The global epidemic and mobility computational model. *Journal of Computational Science*, 1(3):132 – 145, 2010.

[10] Ben S Cooper, Richard J Pitman, W John Edmunds, and Nigel J Gay. Delaying the international spread of pandemic influenza. *PLoS Med*, 3(6):e212, 2006.

[11] Julia Shapiro, Natalie E. Dean, Zachary J. Madewell, Yang Yang, M.Elizabeth Halloran, and Ira Longini. Efficacy estimates for various covid-19 vaccines: What we know from the literature and reports. *medRxiv*, 2021.

[12] Kate M Bubar, Stephen M Kissler, Marc Lipsitch, Sarah Cobey, Yonatan Grad, and Daniel B Larremore. Model-informed covid-19 vaccine prioritization strategies by age and serostatus. *medRxiv*, 2020.

[19] Baisheng Li, Aiping Deng, Kuibiao Li, Yao Hu, Zhencui Li, Qianling Xiong, Zhe Liu, Qianfang Guo, Lirong Zou, Huan Zhang, Meng Zhang, Fangzhu Ouyang, Juan Su, Wenzhe Su, Jing Xu, Huifang Lin, Jing Sun, Jinju Peng, Huiming Jiang, Pingping Zhou, Ting Hu, Min Luo, Yingtao Zhang, Huanying Zheng, Jianpeng Xiao, Tao Liu, Rongfei Che, Hanri Zeng, Zhonghua Zheng, Yushi Huang, Jianxiang Yu, Lina Yi, Jie Wu, Jingdiao Chen, Haojie Zhong, Xiaoling Deng, Min Kang, Oliver G. Pybus, Matthew Hall, Katrina A. Lythgoe, Yan Li, Jun Yuan, Jianfeng He, and Jing Lu. Viral infection and transmission in a large, well-traced outbreak caused by the sars-cov-2 delta variant. *medRxiv*, 2021.

[20] Amanda Minter and Renata Retkute. Approximate bayesian computation for infectious disease modelling. *Epidemics*, 29:100368, 2019.

[21] Mikael Sunnåker, Alberto Giovanni Busetto, Elina Numminen, Jukka Corander, Matthieu Foll, and Christophe Dessimoz. Approximate bayesian computation. *PLOS Computational Biology*, 9(1):1–10, 01 2013.

[22] Centers for Disease Control and Prevention, COVID-19 Pandemic Planning Scenarios. `https://www.cdc.gov/coronavirus/2019-ncov/hcp/planning-scenarios.html#table-1`, 2021. Accessed 2021/02/02.

[23] Google LLC "Google COVID-19 Community Mobility Reports". `https://www.google.com/covid19/mobility/`, 2020. Accessed: 2021-08-01.

[24] Pauli Virtanen, Ralf Gommers, Travis E. Oliphant, Matt Haberland, Tyler Reddy, David Cournapeau, Evgeni Burovski, Pearu Peterson, Warren Weckesser, Jonathan Bright, Stéfan J. van der Walt, Matthew Brett, Joshua Wilson, K. Jarrod Millman, Nikolay Mayorov, Andrew R. J. Nelson, Eric Jones, Robert Kern, Eric Larson, C J Carey, İlhan Polat, Yu Feng, Eric W. Moore, Jake VanderPlas, Denis Laxalde, Josef Perktold, Robert Cimrman, Ian Henriksen, E. A. Quintero, Charles R. Harris, Anne M. Archibald, Antônio H. Ribeiro, Fabian Pedregosa, Paul van Mulbregt, and SciPy 1.0 Contributors. SciPy 1.0: Fundamental Algorithms for Scientific Computing in Python. *Nature Methods*, 17:261–272, 2020.

[25] Nicolò Gozzi, Matteo Chinazzi, Jessica T. Davis, Kunpeng Mu, Ana Pastore y Piontti, Marco Ajelli, Nicola Perra, and Alessandro Vespignani. Anatomy of the first six months of covid-19 vaccination campaign in italy. *medRxiv*, 2021.

[26] Valentina Marziano, Giorgio Guzzetta, Alessia Mammone, Flavia Riccardo, Piero Poletti, Filippo Trentini, Mattia Manica, Andrea Siddu, Antonino Bella, Paola Stefanelli, et al. The effect of covid-19 vaccination in italy and perspectives for living with the virus. *Nature communications*, 12(1):1–8, 2021.
